# A Prospective, Cross-Sectional, Multicenter Pilot Study to Assess the Efficacy of NerveVue: A Non-Invasive, Multi-Site Diagnostic Tool for Peripheral Artery Disease

**DOI:** 10.64898/2025.12.10.25341976

**Authors:** Rakesh Chaitanya, Anuhya Choda, Gayathri Choda

**Affiliations:** Department of Vascular Surgery, The Ramaiah Medical College Hospital, India; Department of Vascular Surgery, Manipal Hospital, India; Aarca Research India Pvt Ltd, Bengaluru, Karnataka, India

**Keywords:** Peripheral Artery Disease (PAD), Ankle Brachial Index (ABI), Non-invasive diagnostics, Vascular screening, PAD classification, Cardiovascular risk screening, Point-of-care device

## Abstract

Peripheral Artery Disease (PAD) remains significantly underdiagnosed even among high-risk populations particularly in primary care and underserved settings due to the dependency on Doppler-based Ankle-Brachial Index (ABI) measurements, which is time-consuming, need a trained personnel and specialized equipment. NerveVue is a novel, portable diagnostic system that integrates photoplethysmography (PPG) and piezoelectric sensors to capture arterial pressure wave form signals from all four limbs simultaneously. Designed for use by low-skill operators, NerveVue enables rapid, infrastructure-light PAD screening suitable for decentralized and outreach environments.

**Methods:** A prospective, cross-sectional, multicenter clinical study was conducted involving 299 adults across standard middleage to older-age ranges. Ankle-Brachial Index (ABI) values derived from the NerveVue device (ABI_NV) were compared against the reference Handheld Doppler method (ABI_Ref). Diagnostic performance metrics including sensitivity, specificity, and predictive values were computed. Subgroup analyses were performed for participants with diabetes, hypertension, and smoking history. Agreement between ABI_NV and ABI_Ref was assessed using Pearson correlation coefficients and PAD classification concordance.

**Results:** NerveVue demonstrated an overall accuracy of 91.6%, with a sensitivity of 84.6% and specificity of 94.7% compared to Handheld Doppler-based ABI measurements. The correlation between ABI_NV and ABI_Ref was strong, with Pearson’s r = 0.991 (right limb) and r = 0.992 (left limb), both statistically significant (p < 0.00001). Subgroup analysis showed consistent performance, with enhanced sensitivity among diabetic participants (90%) and perfect sensitivity among smokers (100%). The system enabled rapid PAD assessment within five minutes using low-skill operators, making it suitable for decentralized screening environments.

**Conclusion:** NerveVue provides
a reliable and accurate alternative to Dopplerbased ABI for the detection of Peripheral Artery Disease (PAD), demonstrating high diagnostic agreement and robust performance across diverse risk groups. Its simultaneous multi-site measurement and ease of use by low-skill operators, position it well for large-scale deployment in community screenings, primary care, diabetic clinics, and preventive health programs. NerveVue has the potential to bridge current diagnostic gaps and enable earlier intervention in vascular disease management.

## Introduction

Peripheral artery disease (PAD) is a progressive atherosclerotic condition characterized by narrowing or occlusion of arteries supplying the lower extremities. It presents clinically across a spectrum, from asymptomatic disease to intermittent claudication and, in advanced cases, critical limb-threatening ischemia. PAD is a marker of systemic vascular disease and is associated with significantly elevated risk for myocardial infarction, stroke, limb amputation, and all-cause mortality ^[1,4,14]^.

Globally, PAD affects more than 200 million people, with a rising burden particularly in low- and middle-income countries (LMICs)^[3]^. Between 2000 and 2010, the estimated prevalence increased by 22% in high-income countries and by 29% in LMICs ^[3]^. PAD prevalence increases sharply with age, affecting over 15–20% of individuals above 70 years, and is especially common in individuals with diabetes, hypertension, chronic kidney disease, and a history of smoking ^[2,3,11]^.

Despite its clinical importance, PAD remains substantially underdiagnosed. It is estimated that 40–60% of individuals with PAD are asymptomatic, and even among symptomatic patients^[5,6]^, the condition is frequently missed in routine clinical practice. Diagnostic underuse is particularly severe in primary care and underserved settings, where reliance on traditional diagnostic modalities, most notably the ankle-brachial index (ABI), creates operational barriers. Doppler-based ABI, while widely accepted as a non-invasive standard, is time-consuming, typically requiring 15–30 minutes per patient, and is operator-dependent, necessitating trained personnel and specialized equipment^[7,8,9]^. These limitations have hindered broader adoption of PAD screening, particularly in decentralized settings where early identification could significantly alter patient outcomes.

To address these gaps, we developed NerveVue, a portable, non-invasive diagnostic system designed to enable rapid PAD screening without requiring Doppler, pressure cuffs, or ultrasound technician expertise. NerveVue integrates photoplethysmography (PPG) and piezoelectric sensors to acquire arterial waveforms from all four limbs simultaneously. It automates waveform synchronization, signal interpretation, and physiologic index generation, enabling minimally trained operators to perform PAD assessments in under three minutes. This study evaluates the diagnostic performance of NerveVue in comparison to Doppler-based ABI in a high-risk, community-based population.

### Device Description

NerveVue is a non-invasive vascular screening system developed by Aarca Research for the early detection and monitoring of Peripheral Artery Disease (PAD). The system is built for rapid, technician-light deployment in clinical and field settings, enabling accessible and scalable vascular risk assessment. The device integrates two complementary sensor modalities, photoplethysmography (PPG) and piezoelectric sensors, into a unified platform designed to capture arterial waveforms from all four limbs simultaneously. PPG sensors detect blood volume changes in peripheral tissues by measuring variations in light absorption related to pulsatile flow. In the context of PAD, PPG is particularly useful for identifying perfusion delays, waveform dampening, and asymmetries in pulse amplitude between upper and lower extremities, features that typically become attenuated in the presence of arterial stenosis or occlusion, thereby serving as non-invasive surrogates for pressure-based perfusion assessment. However, PPG is sensitive to ambient light and individual skin optical properties, which can impact signal quality. To address this limitation, NerveVue incorporates piezoelectric sensors that detect mechanical vibrations associated with arterial wall movement. When placed over superficial arteries such as the radial, dorsalis pedis, or posterior tibial arteries, piezo sensors provide high-fidelity recordings of waveform morphology, capturing both the timing and shape of pressure wave transmitted through the arterial walls. Unlike PPG, this mechanical signal is less affected by skin tone or peripheral perfusion, offering added robustness across diverse populations, including those with diabetes, where peripheral flow may be reduced despite intact arterial mechanics. By fusing waveform data from two sensor types at each limb, NerveVue enables the computation of derived vascular metrics such as Pulse Transit Time (PTT), Pulse Wave Velocity (PWV), amplitude-based ratios, and compliance indicators. These parameters are synthesized into a composite, ABI-informed index (ABI_NV)^[7]^, which enables comparative assessment of limb perfusion symmetry and relative vascular characteristics, including arterial stiffness and compliance.

Unlike traditional ABI, which relies on cuff inflation and Handheld Doppler-guided velocity measurement, NerveVue does not require limb occlusion or skilled probe placement, thus eliminating key barriers to routine PAD screening, particularly in patients with non-compressible arteries or in environments lacking ultrasound trained technicians. The system comprises four sensor modules: clip-on PPG sensors placed on the index fingers and great toes, and clamp-based piezoelectric sensors positioned over the radial and dorsalis pedis or posterior tibial arteries. Arterial waveforms are acquired in parallel for 60 seconds via a microcontroller-based data acquisition unit and transmitted to a companion application via USB or Bluetooth. The software automates waveform analysis, computes ABI_NV, and generates a structured report within 60 sec. Sensor data collection is performed with the participant in a supine position to standardize vascular signal acquisition and minimize posture-related variability. The full screening workflow, from sensor placement to report generation, typically takes less than five minutes. The system is lightweight, portable, and requires minimal operator training. By enabling simultaneous multi-site waveform analysis through a compact low-complexity hardware platform, NerveVue addresses the core limitations of Doppler-based ABI and expands vascular diagnostic capabilities to underdiagnosed and underserved populations

## Materials and Methods

### Study Design and Setting

A prospective, cross-sectional, multicenter clinical validation study was conducted at two community health screening centers affiliated with outpatient clinics of the participating investigators in south India. The primary objective of the study was to evaluate the diagnostic performance of the NerveVue device for the detection of Peripheral Artery Disease (PAD), using Handheld Doppler-based ankle– brachial index (ABI) as the reference gold standard.^[7–9]^

The study was approved by the ACE Independent Ethics Committee (Study Code: NV-2025-01), and was conducted in accordance with the Declaration of Helsinki, the Indian Council of Medical Research (ICMR) National Ethical Guidelines, and ISO 14155 standards for clinical investigation of medical devices. The study was prospectively registered with the Clinical Trials Registry of India (CTRI/2025/11/098233). Written informed consent was obtained from all participants prior to enrolment.

### Participants

A total of 299 participants were enrolled based on the inclusion and exclusion criteria. Recruitment was conducted through voluntary participation. These included routine clinic visits as well as community-based screening activities organized in collaboration with out-patient facility. Demographic information and clinical history, were collected using CRFs and interviews. All measurements were performed in a single session, with participants at rest in the supine position during both reference and investigational device testing.

Each participant underwent testing with the NerveVue device, which simultaneously recorded arterial waveforms from all four limbs using clip-on PPG sensors and clamp-based piezoelectric sensors placed on the radial and dorsalis pedis/posterior tibial arteries For study consistency, all measurements were performed by trained personnel, although the NerveVue system is designed for use by minimally trained operators in routine clinical settings. The complete waveform acquisition took less than 90 seconds per subject. ABI values derived from NerveVue (ABI_NV) were computed through an proprietary algorithm combining pulse transit time (PTT), waveform amplitude symmetry, and morphological pulse features. For reference comparison, all participants underwent standard Ankle-Brachial Index (ABI) measurement using a handheld 5Hz Doppler ultrasound probe and a manual sphygmomanometer. Systolic blood pressures were recorded at the brachial and ankle arteries bilaterally, and ABI_Ref was calculated as the ratio of the higher ankle pressure to the higher brachial pressure. The comparison was performed to evaluate diagnostic agreement, correlation, and performance consistency between the two methods in real-world outpatient and screening settings. PAD was considered present if ABI_Ref was ≤0.90 in either limb. Severity classification of PAD was conducted using standard ABI thresholds^[7,10]^ for both the reference (ABI_Ref) and investigational (ABI_NV) methods. The categories were defined as follows: >1.30 indicating non-compressible arteries, 1.00–1.30 as normal, 0.91–0.99 as borderline, 0.41–0.90 as mild to moderate PAD, and ≤0.40 as severe PAD. These thresholds were used to assign each limb a categorical PAD severity grade, and agreement between ABI_NV and ABI_Ref classifications was analyzed to evaluate category-level concordance.

### Statistical Analysis

The diagnostic performance of ABI values obtained through the NerveVue device (ABI_NV) was evaluated against the reference Doppler method (ABI_Ref) using standard diagnostic metrics: sensitivity, specificity, accuracy, positive predictive value (PPV), and negative predictive value (NPV). Agreement between ABI_NV and ABI_Ref was further assessed using Pearson correlation coefficients (r), with statistical significance determined at p < 0.05. Subgroup analyses were conducted for individuals with diabetes, hypertension, and a history of smoking to evaluate consistency of performance across clinically relevant populations. All analyses were performed using Python (v3.10) and Microsoft Excel.

## Results

### Participant Characteristics

A total of 299 participants were enrolled in the study. The mean age was 48.9 ± 8.7 years, with a gender distribution of 184 males (61.5%) and 115 females (38.5%). The sample included 91 individuals with diabetes (30.4%), 80 with hypertension (26.8%), and a small subgroup reported a history of smoking.smoking (4.7%). All participants underwent both NerveVue and Handheld Doppler ABI measurements in single session.

Based on Doppler ABI (ABI_Ref), 103 participants (34.4%) were classified as having mild to moderate PAD, 75 participants (25.1%) as borderline, and 121 participants (40.5%) had normal ABI values. No cases of severe PAD or non-compressible arteries were identified. This distribution reflects a population at moderate vascular risk, representative of real-world outpatient and community screening environments.

### Overall Diagnostic Performance

ABI values obtained from NerveVue (ABI_NV) demonstrated strong diagnostic agreement with the Doppler-derived ABI_Ref. The overall diagnostic metrics were as follows: sensitivity 84.6%, specificity 94.7%, accuracy 91.6%, positive predictive value 87.5%, and negative predictive value 93.4%. These results reflect high precision in distinguishing both PAD-positive and PAD-negative individuals when benchmarked against the gold-standard method.

### Correlation Between ABI_NV and ABI_Ref

Pearson correlation analysis demonstrated excellent linear agreement between the ABI values derived from the NerveVue device (ABI_NV) and those obtained via the Doppler reference method (ABI_Ref). For the right limb, the correlation coefficient was r = 0.991 (p < 0.00001), and for the left limb, r = 0.992 (p < 0.00001). These results indicate a high degree of measurement concordance between the two methods and support the reliability of waveform-based ABI estimation for PAD.

### Subgroup Performance Analysis

Subgroup analyses demonstrated that NerveVue maintained consistently high diagnostic performance across key vascular risk populations. In participants with diabetes (n = 91), the device achieved a sensitivity of 90.0% and an overall accuracy of 92.9%, with a negative predictive value of 96.8%, highlighting its utility in populations where Doppler ABI may be less reliable. Among hypertensive individuals (n = 80), sensitivity was 86.4%, specificity 96.6%, and overall accuracy 93.8%. In the smoker subgroup (n = 14), NerveVue identified all PAD-positive individuals, achieving 100% sensitivity and 92.9% accuracy. These findings affirm the robustness of NerveVue’s performance across diverse clinical profiles, including groups with known challenges in traditional PAD screening.

## Discussion

This clinical validation study demonstrates that NerveVue, a non-invasive, waveform-based vascular assessment device, delivers diagnostic performance closely aligned with Doppler-based Ankle-Brachial Index (ABI) measurements, the current gold standard for Peripheral Artery Disease (PAD) screening^[7–9]^. While he strong Pearson correlation coefficients (r = 0.991 for the right limb and r = 0.992 for the left limb) underscore the high degree of concordance between NerveVue-derived ABI_NV and the Doppler-derived ABI_Ref, more importantly, these results were obtained using a system that requires minimal technical expertise and completes signal acquisition in under 90 seconds, highlighting NerveVue’s practical ability for decentralized or resource-limited PAD screening environments.^[15,16,17]^

Unlike Doppler-based ABI, which depends on proper cuff inflation, probe handling, and patient rest conditions, NerveVue leverages synchronized piezoelectric and PPG sensors to extract arterial timing and waveform features that are less operator-dependent. While PPG sensors are effective for detecting peripheral perfusion via light absorption changes, they are susceptible to variability from skin pigmentation, ambient light, and perfusion deficits, especially in individuals with diabetes or low cardiac output. The addition of piezoelectric sensors, which measure mechanical pressure waveforms directly from arterial wall pulsations, offers a signal modality that is less affected by skin tone, local perfusion, or ambient conditions. This dual-sensor approach increases robustness and reliability. By capturing both volumetric and mechanical signals, NerveVue improves the precision of pulse transit timing, waveform morphology analysis, and overall signal confidence-enhancing PAD detection accuracy across a broader patient population.

Subgroup analysis further reinforces the system’s robustness across clinically significant populations. In individuals with diabetes, a group where medial arterial calcification often leads to falsely elevated ABI values^[12,13]^, NerveVue achieved a sensitivity of 90.0% and an overall accuracy of 92.9%. This suggests that waveform-based assessment may overcome the limitations of pressure-based diagnostics in detecting perfusion abnormalities in calcified vessels. Similarly, strong performance was maintained in hypertensive individuals (accuracy 93.8%) and smokers (100% sensitivity), two groups with elevated PAD risk and variable vascular compliance. These findings are particularly relevant for settings where diverse patient profiles and comorbidities are common, and where conventional ABI may have known reliability issues.

Taken together, the data support the role of NerveVue as a viable alternative to Doppler ABI for PAD screening. Its rapid, non-invasive, and low-training operation offers a pathway for integration into high-throughput screening programs, opportunistic assessments during outpatient visits, or chronic disease management workflows. Moreover, the system’s digital reporting and connectivity infrastructure make it amenable to telemedicine and population health applications. Future studies with larger and more diverse cohorts— including participants with severe PAD and non-compressible arteries—are warranted to further characterize the system’s diagnostic boundaries. Longitudinal studies assessing predictive outcomes and cost-effectiveness in clinical practice would also strengthen the case for adoption in national screening strategies.

### Limitations

he study did not include patients with compressible arteries, which may critical limb ischemia or those with non-influence diagnostic accuracy. Additionally, longitudinal outcomes following NerveVue-based diagnosis were not assessed. Future studies may explore device performance in follow-up monitoring and its ability to detect vascular improvement or progression.

## Conclusion

This study demonstrates that NerveVue—a portable, non-invasive vascular diagnostic system—provides a reliable and scalable alternative to Doppler-based Ankle-Brachial Index (ABI) measurement for Peripheral Artery Disease (PAD) screening. With high sensitivity, specificity, and strong correlation to the reference standard, NerveVue shows consistent diagnostic performance even in high-risk populations where traditional ABI may be limited, such as individuals with diabetes or hypertension. By enabling rapid, technician-light, multi-site waveform acquisition and automated analysis, the device addresses key operational challenges that currently hinder widespread PAD detection in primary care and community-based settings. Its integration into routine screening workflows has the potential to expand vascular health access, support timely referrals, and reduce the burden of undiagnosed PAD^[21,22]^. Further research will focus on longitudinal follow-up, real-world implementation in high-throughput clinics, and ongoing algorithmic development to support broader vascular profiling and risk stratification.

**Figure 1:**
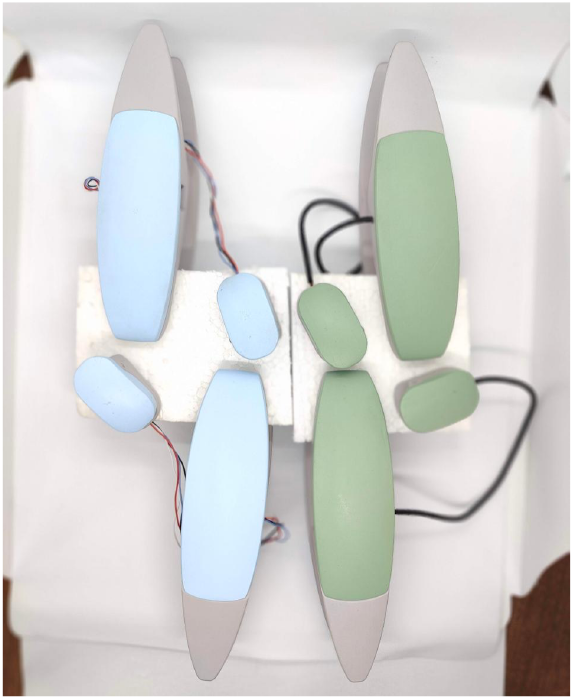
NerveVue Device.

**Figure 2:**
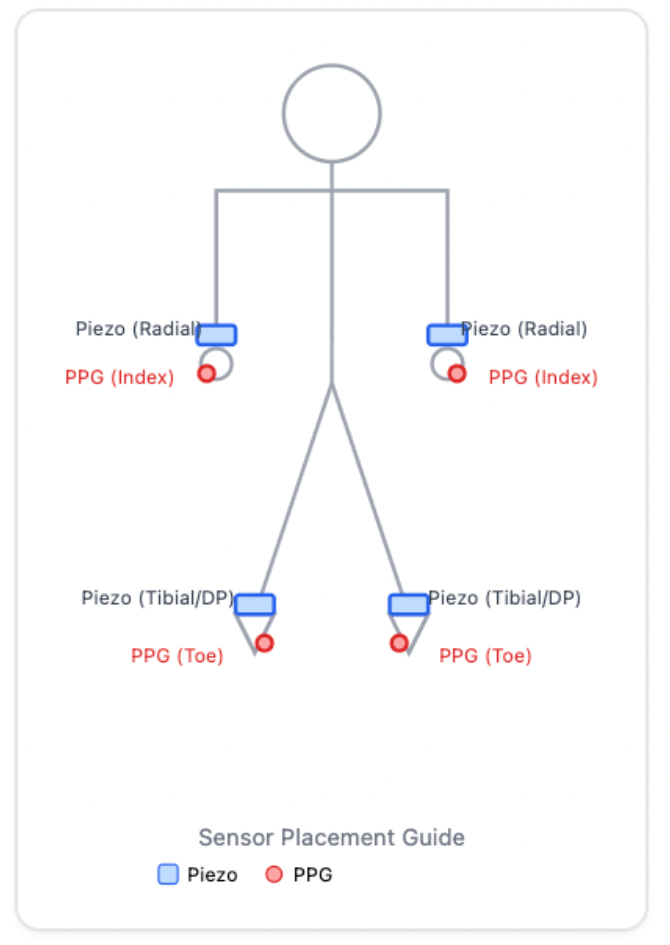
NerveVue sensor placement.

**Figure 3:**
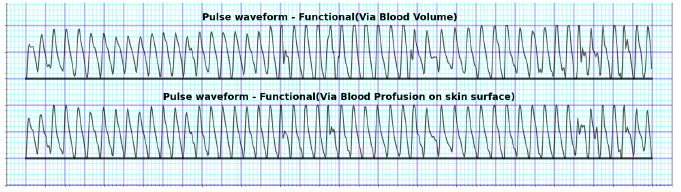
NerveVue Waveforms.

**Table 1:**
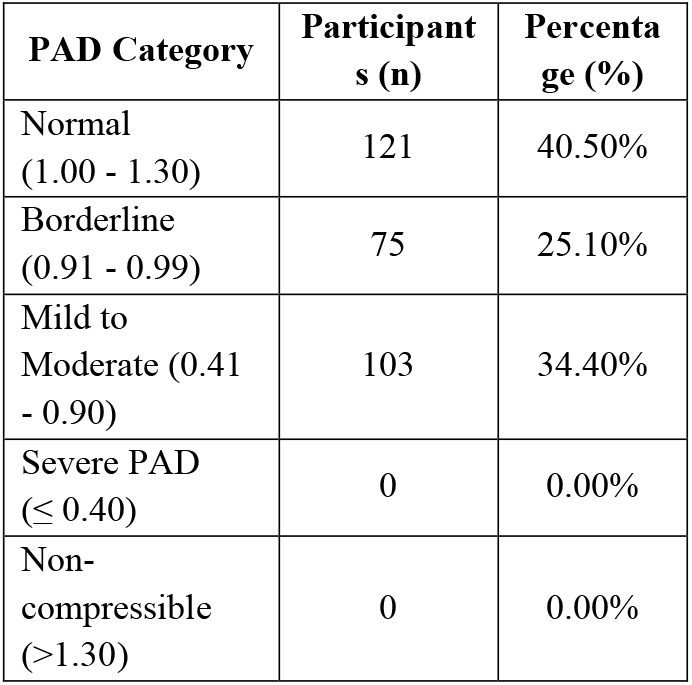
Participant Characteristics1.

**Figure 3: NerveVue Waveforms**
| Table 1: Participant Characteristics1 |  |  |
| --- | --- | --- |
| PAD Category | Participant<br>s (n) | Percenta<br>ge (%) |
| Normal<br>(1.00 - 1.30) | 121 | 40.50% |
| Borderline<br>(0.91 - 0.99) | 75 | 25.10% |
| Mild to<br>Moderate (0.41<br>- 0.90) | 103 | 34.40% |
| Severe PAD<br>(≤ 0.40) | 0 | 0.00% |
| Non-compressible<br>(>1.30) | 0 | 0.00% |

**Table 2:**
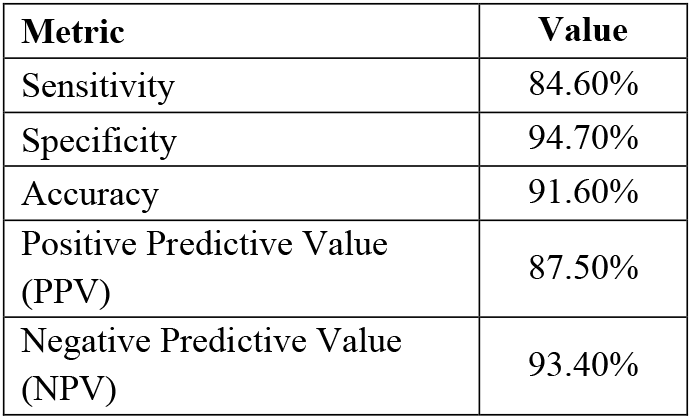
PAD Classification by Doppler ABI.

**Table 3:** Overall Diagnostic Performance of NerveVue.

| Table 3: Overall Diagnostic Performance of NerveVue |  |  |
| --- | --- | --- |
| Limb | Pearson<br>Correlation<br>Coefficient<br>(r) | p-value |
| Right | 0.991 | < 0.00001 |
| Left | 0.992 | < 0.00001 |

**Table 4:** Correlation Between ABI_NV and ABI_Ref.

| Table 4: Correlation Between ABI_NV and ABI_Ref |  |
| --- | --- |
| Metric | Value |
| Sensitivity | 84.60% |
| Specificity | 94.70% |
| Accuracy | 91.60% |
| Positive Predictive Value<br>(PPV) | 87.50% |
| Negative Predictive Value<br>(NPV) | 93.40% |

**Table 5:** Subgroup Performance of NerveVue.

| Table 5: Subgroup Performance of NerveVue |  |  |  |
| --- | --- | --- | --- |
| Metric | Diabet<br>es<br>(n =<br>91) | Hypertensi<br>on (n = 80) | Smoki<br>ng<br>history<br>(n =<br>14) |
| Sensitivit<br>y | 90.00<br>% | 86.40% | 100% |
| Specificit<br>y | 0 | 96.60% | 0 |
| Accuracy | 92.90<br>% | 93.80% | 92.90% |
| PPV |  |  |  |
| NPV | 96.80<br>% |  |  |

## Data Availability

All data produced in the present study are available upon reasonable request to the authors.

## Acknowledgements

The authors thank the clinical and field staff who supported patient screening and data collection. We acknowledge the participants for their time and consent. This study was conducted under CDSCO Test License. Funding support was provided in part by the Pfizer Indovation Grant for Aarca Research India Pvt Ltd.

## Author Contributions

Rakesh Chaitanya served as the Principal Investigator and provided overall clinical oversight, patient evaluation, and protocol development, supervision of screening procedures, and data review. Gayathri Choda led the study design, device integration and workflow development, coordination of data acquisition, and manuscript preparation. All authors reviewed and approved the final manuscript.

